# Artificial Intelligence Assisted Early Warning System for Acute Kidney Injury Driven by Multi-Center ICU Database

**DOI:** 10.1101/2020.01.27.20019091

**Authors:** Sai Huang, Li Chen, Lili Wang, Xiang Cui, Cong Feng, Zhengbo Zhang, Tanshi Li

## Abstract

**Background:** To improve the performance of early acute kidney injury (AKI) prediction in intensive care unit (ICU), we developed and externally validated machine learning algorithms in two large ICU databases.

**Methods:** Using eICU^®^ Collaborative Research Database (eICU) and MIMIC-III databases, we selected all adult patients (age ≥ 18). The detection of AKI was based on both the oliguric and serum creatinine criteria of the KDIGO (Kidney Disease Improving Global Outcomes). We developed an early warning system for forecasting the onset of AKI within the first week of ICU stay, by using 6- or 12-hours as the data extraction window and make a prediction within a 1-hour window after a gap window of 6- or 12-hours. We used 52 features which are routinely available ICU data as predictors. eICU was used for model development, and MIMIC-III was used for externally validation. We applied and experimented on eight machine learning algorithms for the prediction task.

**Results:** 3,816 unique admissions in multi-center eICU database were selected for model development, and 5,975 unique admissions in single-center MIMIC-III database were selected for external validation. The incidence of AKI within the first week of ICU stay in eICU and MIMIC-III cohorts was 52.1% (n=1,988) and 31.3% (n=1,870), respectively. In eICU cohort, the performance of AKI prediction is better with shorter extraction window and gap window. We found that the AdaBoost algorithm yielded the highest AUC (0.8859) on the model with 6-hours data extraction window and 6-hours gap window (model 6-6) rather than other prediction models. In MIMIC-III cohort, AdaBoost also performed well.

**Conclusions:** We developed the machine learning-based early AKI prediction model, which considered clinical important features and has been validated in two datasets.

## Background

Acute kidney injury (AKI) is the major case of multiple organ failure with an incidence more than 50% in critically ill patients ^1-5^. Early prevention as one of the limited management options could greatly improve the prognosis of AKI in the intensive care unit (ICU) and therefore the early identification of critical patients with AKI becomes a critical issue, which could offer an opportunity to develop strategies for early prevention and intervention of AKI in critical care ^6-9^. The available dataset of large electronic healthcare records (EHR) provide an opportunity of predicting the early onset of AKI in critical care patients using machine learning algorithms, which can provide a much earlier, applicable and cost-effective solution than current late diagnostic criteria and biomarkers with limited utilities ^6, 10-18^.

Forecasting the onset of AKI was well-studied in previous. Some studies have reported that their AKI prediction models were desirable. However, deploying such methods to the real world would be relatively difficult due to several critical limitations ^19-21^: (1) Some prediction models were based on highly specific cohorts, such as cardiac disease and post-surgery cohorts, which lack of the general hospital population background, thus need to be further explored ^22-27^. (2) The definitions of AKI in some studies were based on the serum creatinine (SCr) criteria without oliguria criteria. Even though oliguria criteria is non-early and non-specific, some studies have already confirmed that nearly one-third of AKI patients in ICU were identified by oliguria criteria and without elevating SCr^26-36^. Some of the previous study such as Tomašev *et al*. 2019 were due to the lack of urine output information. (3) Those prediction tasks simply rely on a single time point, which is the time of admission to hospital, due to data insufficiency ^26, 27, 31, 32, 37-40^. (4) Many studies did not include physiological or laboratory parameters as predictors, especially urine output, which has been confirmed as a critical role in predicting AKI and ICU mortality ^26, 27, 34, 35, 41^. (5) The explored machine learning methods and selected features in the studies were limited, which may not fully take the advantage of machine learning techniques based on “big healthcare data” as well as investigating the best model performance ^26, 31-36, 39, 40, 42^. For example, Tomašev *et al*. 2019 developed a deep learning approach for the continuous risk prediction of AKI, but they did not compare their approach with other baseline algorithms such as basic neural networks or other ensemble algorithms. (6) Many studies did not perform external validation or not considering the real-world deployment ^26, 31-36, 40, 43-47^.

To overcome the above limitations, we utilized a large multi-center ICU database (eICU Collaborative Research Database (v1.2)) and used both oliguria and SCr diagnostic criteria of Kidney Disease Improving Global Outcomes (KDIGO) ^14^ as the definition of AKI, and included most of the routinely available physiological and laboratory parameters in ICU and clinical interventions as predictors. We then adopted eight machine learning algorithms to conduct experiments and develop the comparable models of predicting the early onset of AKI within the first week of ICU stay. We also used another large non-overlapping single-center MIMIC-III (Medical Information Mart for Intensive Care III (v1.4)) ICU database for external validation. By fully utilizing the existed databases, we evaluated four time series models in an artificial intelligence perspective. The most important clinical features are also identified by feature importance analysis.

## Methods

### Study Design and Databases

We developed an early warning model for prediction the onset of AKI based on the retrospective analysis of the multi-center eICU database. The database including 459 ICUs from 58 hospitals during 2003–2016 in the United States ^48, 49^. The model was externally validated on a single-center publicly available ICU database (MIMIC-III database), which contains 5 different ICUs in a tertiary medical center (Beth Israel Deaconess Medical Center, BIDMC; Boston, Massachusetts) during 2001–2012 ^50, 51^.

### Study Cohorts

All patients in the publically available eICU (v1.2) and MIMIC-III (v1.4) databases were considered in this study. The inclusion criteria are as follows: 1) Age ≥ 18 years old ^52^; 2) ICU stay for at least 24 consecutive hours; and 3) The first ICU admission of the first hospitalization were used ^53^. The exclusion criteria are as follows: 1) The primary diagnosis contained “Kidney disease”, “Acute renal failure”, and “Renal obstruction”; 2) clinical history of end-stage renal disease (ESRD) or their SCr was ≥4 mg/dl within the first 24 hours of ICU admission ^26^; 3) Received renal replacement therapy (RRT) within the first 24 hours of ICU admission ^54^; 4) diagnosed as AKI based on oliguria diagnostic criteria within the first 24 hours of ICU admission ^54, 55^; 5) Withdrawal of treatment (which contains patients who did not receive mechanical ventilation and died during ICU stay); 6) samples with any missing value after imputation ^53^.

### AKI Definition

The detection of AKI in adults was based on the oliguric and SCr criteria of the KDIGO ^56^. The “baseline” of SCr was defined as the ICU admission. ^32, 37, 38, 57, 58^. The oliguric criteria is based on the urine output <0.5 ml/kg/h for more than 6 hours or 12 hours, or the urine output <0.3 ml/kg/h for 24 hours ^56^.

### Prediction Task and Outcomes

The prediction target or the primary outcome of this study was the first onset of AKI at any stage within the first week of ICU stay ^31, 52, 53^. We used 6 or 12 hours ^54, 55, 59^ as the feature collecting window, since the urine output based diagnostic criteria of KDIGO relies on 6 or 12 consecutive hours, and we make a one-hour prediction window after a gap window of 6 or 12 hours. That is, we used 6 or 12 hours of data to forecast the onset of AKI after 6 or 12 hours. All features and labels are extracted at one-hour interval for time-series modeling (Figure 1). Although sampled in one-hour, forecasting the future onset of AKI is a continuous prediction over the time.

**Figure 1.**
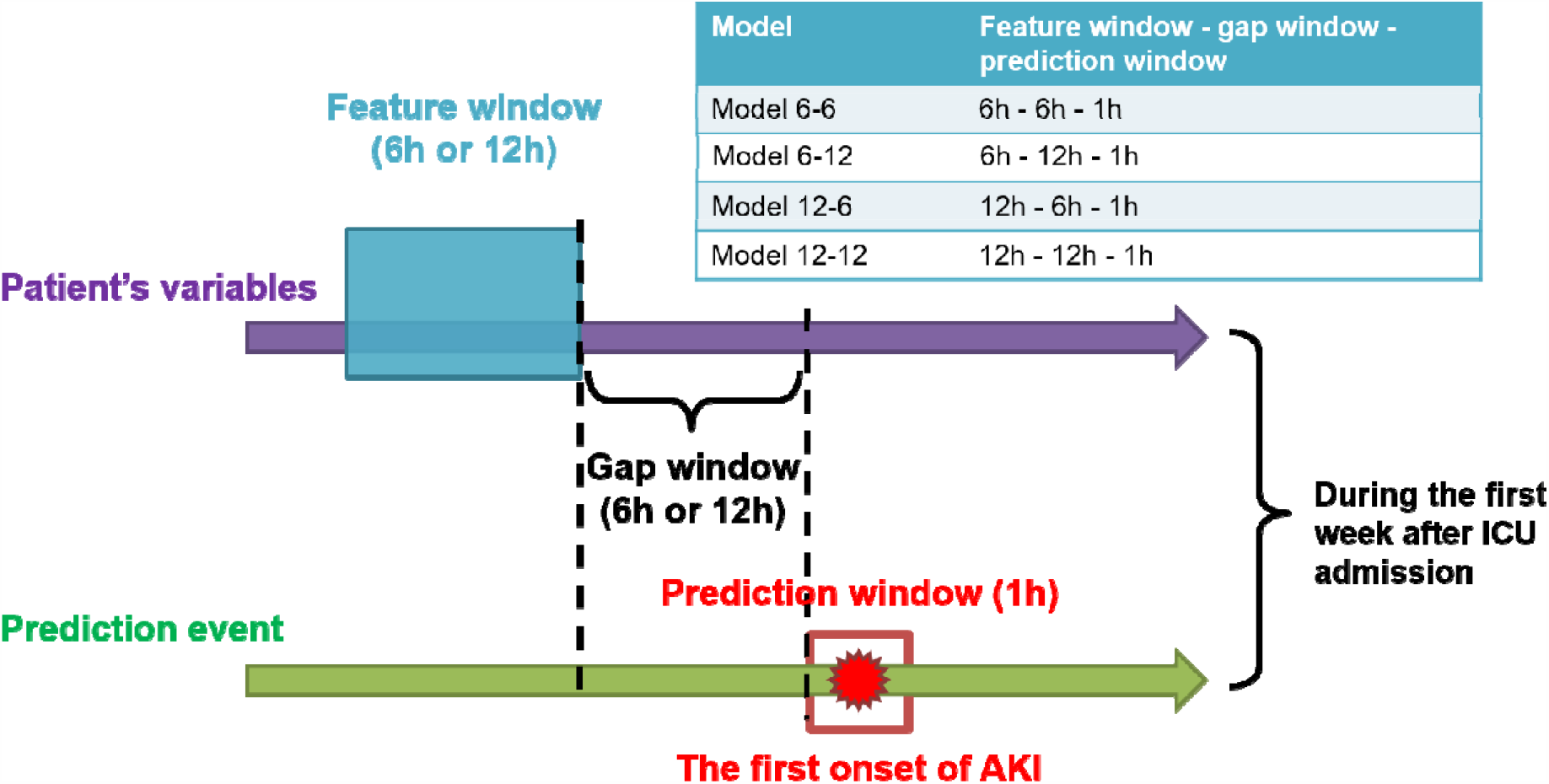
Model settings and prediction for a single admission. Timestamp is at one-hour interval. Feature collecting window are the discretized time-series features (6 hours or 12 hours) as inputs of machine learning algorithms for predicting the label of AKI after certain gaps (6 hours or 12 hours). We experimented on four different model configurations: model 6-6, model 6-12, model 12-6, and model 12-12.

In detail, we experimented four configurations of time-series models. The configurations are described as follow with model name and the hours of feature collecting window – the hours of gap window – the hours of prediction window, respectively.

1. Model 6-6: 6h-6h-1h;
2. Model 6-12: 6h-12h-1h;
3. Model 12-6: 12h-6h-1h;
4. Model 12-12: 12h-12h-1h.

Secondary outcomes of the study included ICU mortality, initial sequential organ failure assessment (ISOFA) score, need for renal replacement therapy (RRT) and ICU length of stay (LOS) ^31, 52, 53^. The prediction task and different models are shown in Figure 2.

**Figure 2.**
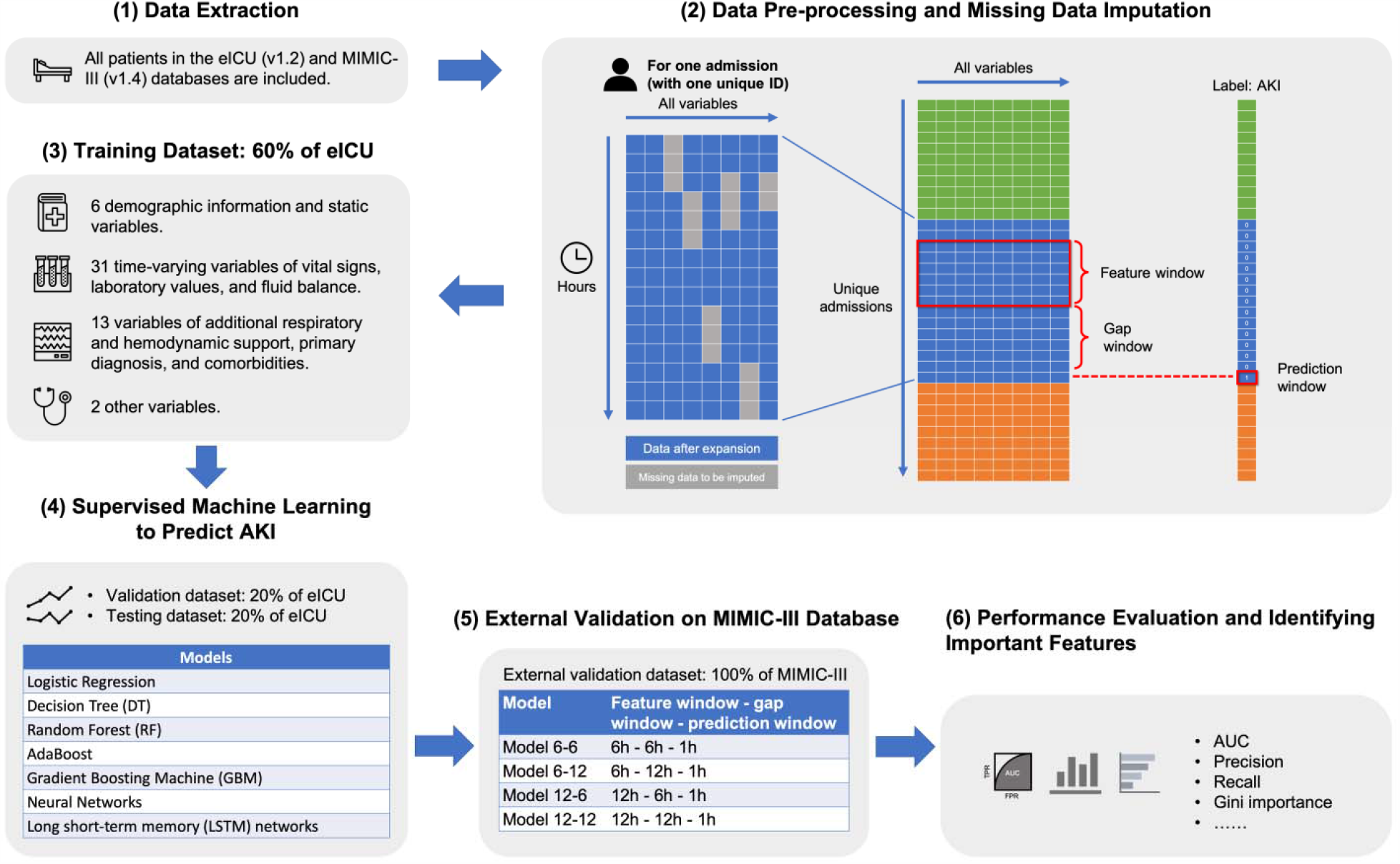
The study overview of the artificial intelligence-assisted AKI early warning system, including (1) Data extraction, (2) Data pre-processing and missing data imputation, (3) dataset splitting, (4) supervised machine learning method to predict AKI, (5) external validation, and (6) performance evaluation and important feature identification.

### Model Comparison and Feature Importance Ranking

The violin plot ^60^ and pairwise paired t-test (two sided) ^61^ were used to visualize and evaluate th performances across four different configurations of our time-series models (model 6-12; model 6-6; model 12-12, and model 12-6). Gini importance ^62^, also known as the mean decrease impurity, is averaged over all trees of the ensemble model, which was used for identifying and ranking feature importance. Gini importance is calculated by scikit-learn packages ^63^.

### Feature Examination and Secondary Outcomes

We used “psych” package ^64^ to further examine the association between ICU mortality, length of stay (LOS), need for renal replacement therapy (RRT), and the occurrence of AKI in the first week to obtain the secondary outcomes, since these features were collected after the onset of AKI and not showed in Table 1. Moreover, ISOFA score was also included into secondary outcomes. Continuous features are shown as the mean with standard deviation (s.d.) or median with interquartile range (IQR). Categorical features are expressed as absolute (n) and relative (%) frequency. Selected features were analyzed by the Spearman *ρ* correlation test (two-sided), where the Spearman *ρ* statistic is used to estimate a rank-based measure of association. The test is a non-parametric and distribution-free test, thus the data do not necessarily follow a bivariate normal distribution ^65^. The significance was considered when P < 0.05. Statistical analysis was performed by Python 3.7 and R version 3.5.1.

**Table 1.**
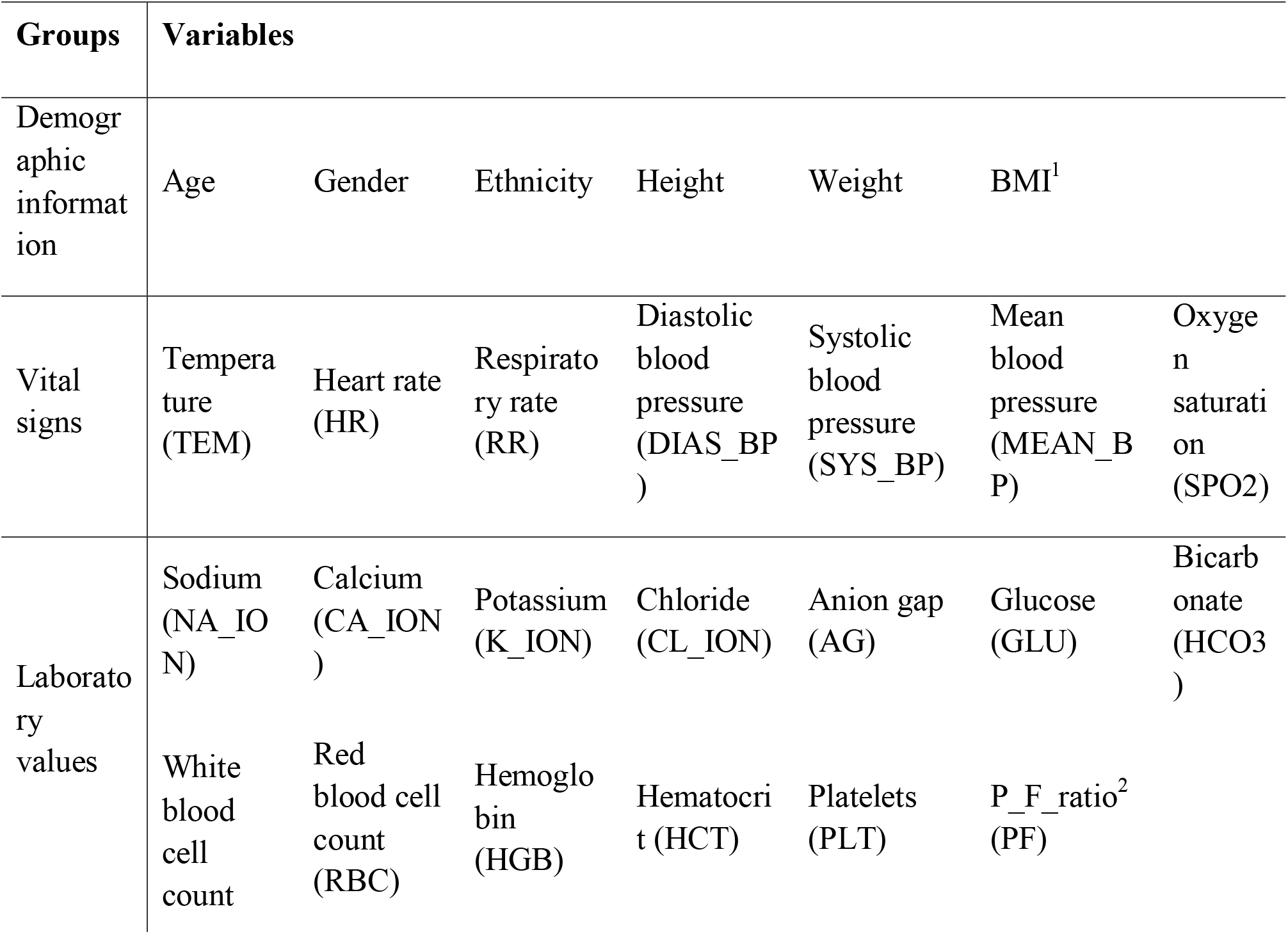
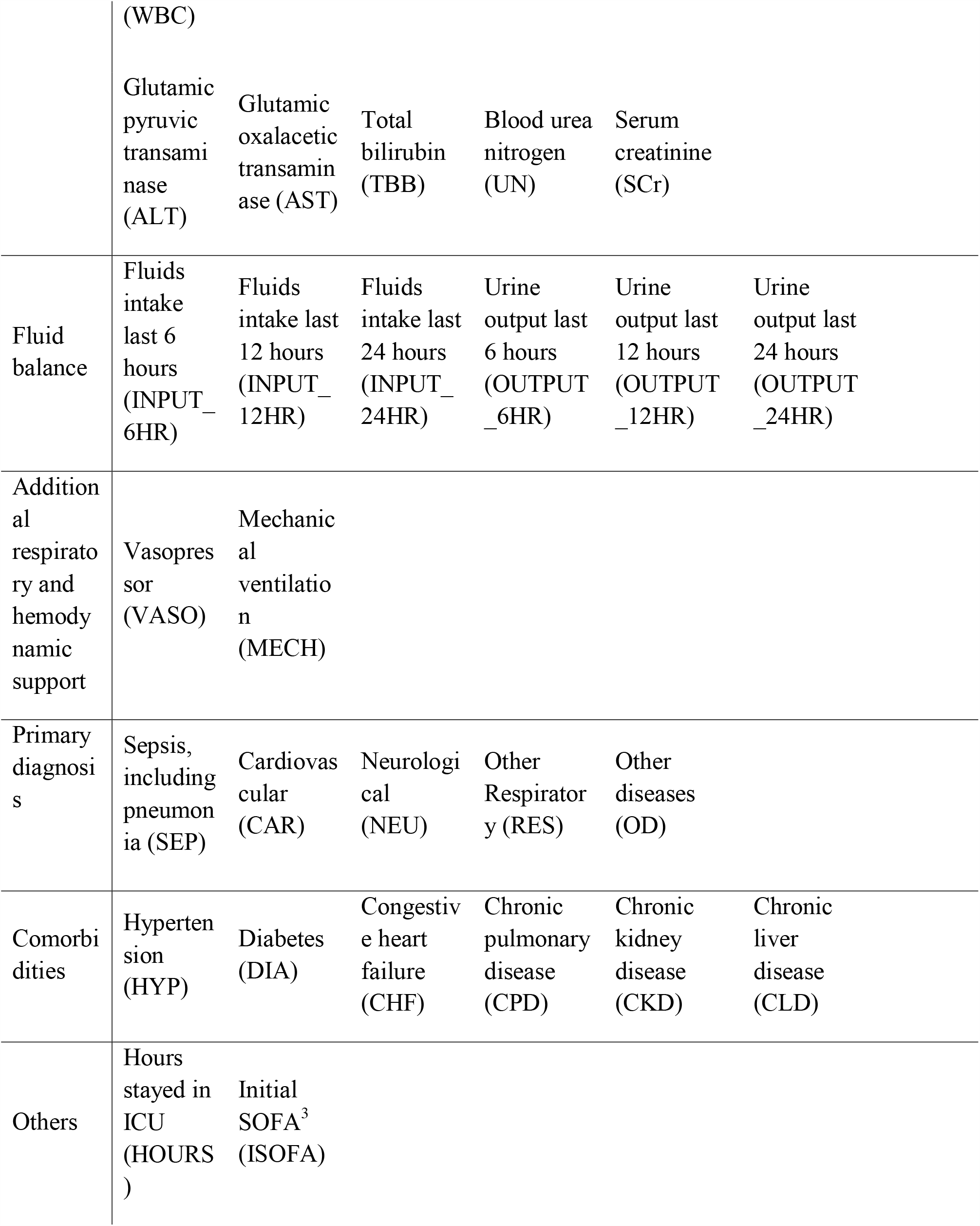
Variables and categorized groups. ^1^Body Mass Index: Weight (kg) / Height (meter) * Height (meter). BMI then converted to classes according to following criteria: BMI < 18.5: class = 0; 18.5 ≤ BMI < 23: class = 1; 23 ≤ BMI < 25: class = 2; 25 ≤ BMI < 40: class = 3; BMI ≥ 40: class = 4. ^2^P_F_ratio: PO2/FiO2. ^3^SOFA: Sequential Organ Failure Assessment.

| Groups | Variables |  |  |  |  |  |  |
| --- | --- | --- | --- | --- | --- | --- | --- |
| Demographic information | Age | Gender | Ethnicity | Height | Weight | BMI<sup>1</sup> |  |
| Vital signs | Temperature (TEM) | Heart rate (HR) | Respiratory rate (RR) | Diastolic blood pressure (DIAS_BP) | Systolic blood pressure (SYS_BP) | Mean blood pressure (MEAN_BP) | Oxygen saturation (SPO2) |
| Laboratory values | Sodium (NA_ION) | Calcium (CA_ION) | Potassium (K_ION) | Chloride (CL_ION) | Anion gap (AG) | Glucose (GLU) | Bicarbonate (HCO3) |
|  | White blood cell count | Red blood cell count (RBC) | Hemoglobin (HGB) | Hematocrit (HCT) | Platelets (PLT) | P_F_ratio<sup>2</sup> (PF) |  |

Table 1. Variables and categorized groups. <sup>1</sup>Body Mass Index: Weight (kg) / Height (meter) \*
|  |  |  |  |  |  |  |
| --- | --- | --- | --- | --- | --- | --- |
|  | (WBC) |  |  |  |  |  |
|  | Glutamic<br>pyruvic<br>transami<br>nase<br>(ALT) | Glutamic<br>oxalacetic<br>transamin<br>ase (AST) | Total<br>bilirubin<br>(TBB) | Blood urea<br>nitrogen<br>(UN) | Serum<br>creatinine<br>(SCr) |  |
| Fluid<br>balance | Fluids<br>intake<br>last 6<br>hours<br>(INPUT_<br>6HR) | Fluids<br>intake last<br>12 hours<br>(INPUT_<br>12HR) | Fluids<br>intake last<br>24 hours<br>(INPUT_<br>24HR) | Urine<br>output last<br>6 hours<br>(OUTPUT<br>_6HR) | Urine<br>output last<br>12 hours<br>(OUTPUT<br>_12HR) | Urine<br>output last<br>24 hours<br>(OUTPUT<br>_24HR) |
| Addition<br>al<br>respirato<br>ry and<br>hemody<br>namic<br>support | Vasopres<br>sor<br>(VASO) | Mechanic<br>al<br>ventilatio<br>n<br>(MECH) |  |  |  |  |
| Primary<br>diagnosi<br>s | Sepsis,<br>including<br>pneumon<br>ia (SEP) | Cardiovas<br>cular<br>(CAR) | Neurologi<br>cal<br>(NEU) | Other<br>Respirator<br>y (RES) | Other<br>diseases<br>(OD) |  |
| Comorbi<br>dities | Hyperten<br>sion<br>(HYP) | Diabetes<br>(DIA) | Congestiv<br>e heart<br>failure<br>(CHF) | Chronic<br>pulmonary<br>disease<br>(CPD) | Chronic<br>kidney<br>disease<br>(CKD) | Chronic<br>liver<br>disease<br>(CLD) |
| Others | Hours<br>stayed in<br>ICU<br>(HOURS<br>) | Initial<br>SOFA<sup>3</sup><br>(ISOFA) |  |  |  |  |

### Data Extraction and Pre-processing

#### Predictors

For each patient, we extracted most of the routinely available ICU data as predictors based on methods of previous studies ^31, 42, 54, 59, 66^ and the availability in the selected databases. We included following features: 1) 18 static variables of demographic information, primary diagnosis and comorbidities (*e*.*g*., age, gender, sepsis, hypertension, etc.). The primary diagnosis were identified at the ICU admission from the databases; 2) 31 time-varying variables of vital signs, laboratory values and fluid balance-related parameters (*e*.*g*., heart rate, oxygen saturation, white blood cell count, blood creatinine, fluids intake, urine output, etc.); 3) two critical interventions, which are usages of mechanical ventilation and vasopressors; 4) two other variables: “Hours” (hours stayed in ICU) and Initial SOFA (ISOFA). The complete feature list is shown in Table 1, detailed feature selection criteria are listed in Table S1.

It is worth to know that those variables also include urine output and serum creatinine (SCr), the same two variables which were used to define the onset of AKI based on KDIGO ^56^. In time series analysis, these two variables are known as endogenous variables while other variables are served as exogenous variables ^67, 68^. Thought the definition of AKI is based on urine output and SCr, forecasting the onset of AKI after 6 or 12 hours gap window cannot base on the future unseen urine output and SCr. Thus, complex machine learning algorithms are involved into this forecasting problem.

### Dataset Partitioning and Missing Data Imputation

eICU database was used for model development and the data was analyzed with five-fold cross-validation: *i*.*e*., randomly split the entire cohorts into 60%:20%:20% as training, validation and testing sets, respectively. The training set (60%) is used for developing machine learning models, the validation set (20%) helps algorithms find their best hyper-parameters learned by the training set. The testing set (20%) is used for evaluating the performances of the models learned and optimized by training and validation sets. To prevent bias introduced when different admissions of one patient appears in separate dataset, data was splitted according to their unique patient ID instead of the admission ID, and assigned into either train, validation, or test set, to ensure patients are mutually exclusive across different datasets.

Missing values were imputed with linear imputation approach ^69^ with respect to each admission. Once the linear imputation is done, nearest-neighbor imputation ^70-72^ was adopted within each admission to fill the remained missing values outside of the linear imputation range. All imputations were performed piece-wisely on each single admission. As linear imputation was adopted, it is critical to avoid data leakage on the validation and testing datasets. In our scenario, each variable on each admission in validation and testing sets are either partially missing or complete, otherwise that admission will be dropped. We then let those admissions impute the missing value by itself instead of using the training set. Moreover, the onset of AKI is occurred at the last timestamp of each admission since the data after the onset of AKI or at the time of AKI were not used in model-building, thus, the experiment will not have the data leakage issue on time and will not bias the prediction after the data imputation.

Data were coded as multi-dimensional time-series at one-hour interval ^66^. Static variables (demographic information, primary diagnosis, comorbidities, and ISOFA) were replicated across all timestamps for each patient. Categorical features with multiple categories but without order information, such as “ethnicity”, were one-hot encoded by using “OneHotEncoder” function from scikit-learn package ^63^. Vital signs and laboratory values with multiple measurements within one-hour interval were pooled in average ^66^ and rounded to the nearest unit hour.

### Model Development and Validation

Driven by the large population database, we used eight different machine learning algorithms to develop our early AKI prediction model. The algorithms include: logistic regression (baseline), random forest (RF) ^73^, AdaBoost (short for Adaptive Boosting, base estimator: decision tree with maximum depth = 1) ^74^, decision tree (DT), Gradient Boost Machine (GBM) ^75^, neural network (NN), L2 regularized neural network (NN-L2), and long short-term memory (LSTM) neural networks ^76^. Then we used MIMIC-III database for external validation. Time-series data were encoded and reshaped to 1-D vectors to feed the learning algorithms except for LSTM. Hyper-parameters are fine-tuned for each algorithm on validation set. The entire workflow is shown in Figure 2.

### Performance Evaluation and Statistical Analysis

As logistic regression was widely used in predicting time-series events for ICU-related problems ^26, 54, 66^, we used logistic regression as the baseline for comparisons. Performances was evaluated by: 1) the area under the ROC curve (AUC) ^77^; 2) Precision (macro) ^78^; 3) Recall (macro) ^78^; 4) F1 score (macro) ^79^; 5) negative predictive value ^80^; and 6) specificity ^81^. AUC, ranging from 0 to 1, the higher the better, indicates the algorithm’s performances. We considered the model with AUC above 0.85 as the adequate and well-performed model. Model with the highest AUC is considered as the best model. Precision, also known as positive predictive value (PPV) ^80^, is the fraction of true positive classification among the positive results classified by algorithm, a higher precision indicates an algorithm’s result is reliable. Recall, also known as sensitivity ^81^, is the fraction of true positive classification among all the true samples, describes the ability of identifying true samples (AKI) among the whole population. F1 score is the harmonic average of precision and recall, higher F1 score indicates better performance. As we reported PPV (precision) and sensitivity (recall), we also took negative predictive value (NPV) and specificity into account. NPV is the probability that an admission with a negative prediction truly don’t have the AKI. Specificity measures the proportion of actual negatives samples that are correctly identified. Since five-fold cross-validation training scheme was adopted which has five different mutually exclusive testing sets (each contains 20% of the entire cohort), thus all the performance metrics are computed 5 times. Results are either presented by mean values in Tables or by boxplot in Figures.

## Results

### Study Cohorts: eICU and MIMIC-III

A total of 3,816 unique admissions of eICU database and 5,975 unique admissions of MIMIC-III database were included after data pre-processing and filtering by inclusion and exclusion criteria. Detailed inclusion and exclusion process are presented in Figure 3.

**Figure 3.**
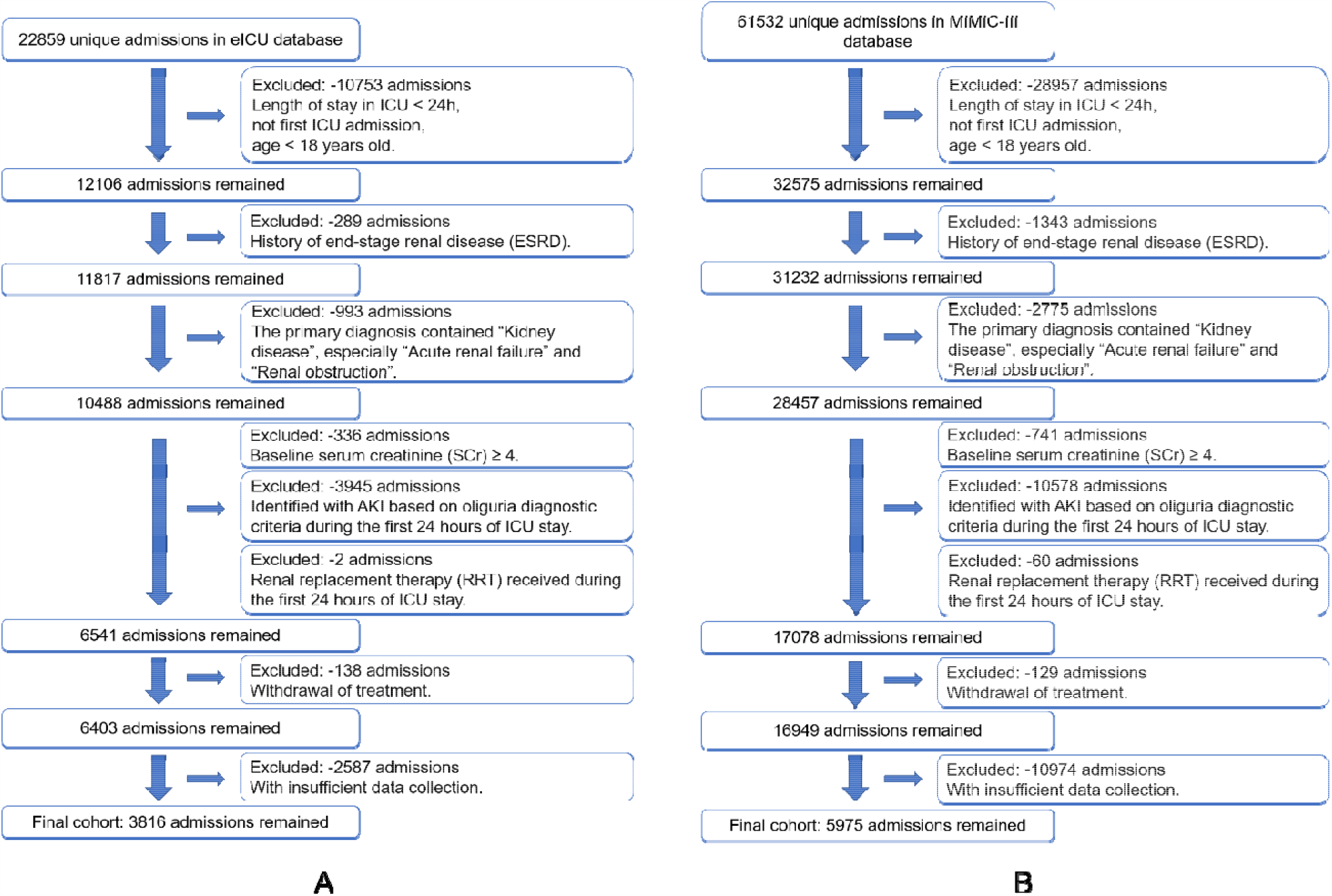
Study cohort selection workflow of eICU (A) and MIMIC-III database (B) based on the designed inclusion and exclusion criteria.

Descriptions and statistics of patients’ demographics and clinical characteristics between two cohorts are summarized in Table 2. Commonly missing features are also described in Table S2. The eICU cohort contained 3,816 unique admissions with a mean age of 60 (standard deviation=18.0); 56.8% (n=2,169) were male and 84.8% (n=3,234) were white (including Eastern European, Brazilian, Russian, and other European). 5,975 unique admissions selected in the final MIMIC-III cohort with a mean age of 60 (standard deviation=18.3); 57.0% (n=3,404) were men and 69.3% (n=4,140) were white. The incidence of AKI within the first week in eICU and MIMIC-III cohorts was 52.1% (n=1,988) and 31.3% (n=1,870), respectively. The AKI cases were determined based on urine output criteria in eICU and MIMIC-III cohorts were 45.1% (n=1,722) and 24.9% (n=1,486). Sepsis, including pneumonia, as the primary diagnosis accounted for 18.7% (n=713) in eICU cohort and 38.39% (n=2,290) in MIMIC-III cohort. Cardiovascular disease accounted for 35.8% (n=1367) in eICU cohort and 25.6% (n=1,531) in MIMIC-III cohort. The proportion of patients on mechanical ventilation and vasopressors were 33.4% (n=1,276) and 26.4% (n=1,006) in the eICU, 47.4% (n=2,830) and 27.9% (n=1,666) in the MIMIC-III. The proportion of patients on cardiac surgery prior to AKI were 3.9% (n=149) in the eICU and 2.0% (n=122) in the MIMIC-III. The mean ISOFA score was 5.8 (standard deviation=3.3) in the eICU and 4.1 (standard deviation=3.0) in the MIMIC-III. The need for renal replacement therapy (RRT) was 1.8% (n=68) in the eICU and 0.95% (n=57) in the MIMIC-III. Median ICU length of stay was 2.6 days in eICU and 2.8 days in MIMIC-III. The proportion of ICU mortality was 10.0% in eICU and 8.6% in MIMIC-III.

**Table 2.** Description of the datasets. Ethnicity of Black includes African American, Cape Verdean, Haitian, and African. Ethnicity of Hispanic includes Latino (central American), Latino (Cuban), Latino (Puerto Rican), Latino (Honduran), Latino (Guatemalan), Latino (Mexican), Latino (Dominican), Latino (Salvadoran), Latino (Colombian), and Portuguese. Ethnicity of Asian includes Vietnamese, Thai, Asian Indian, middle Eastern, Korean, Chinese, Filipino, Cambodian, Japanese, Asian other. White includes Eastern European, Brazilian, Russian, and other European. Ethnicity of Other includes Unknown, not specified, multi race, patient declined to answer, and unable to obtain. s.d.: Standard deviation in short. IQR: interquartile range in short.

| Key Characteristics | eICU | MIMIC-III |
| --- | --- | --- |
| Unique ICU types (n) | 8 | 5 |
| Final cohort (n) | 3,816 | 5,975 |
| The incidence of AKI within the first week (n(%)) | 1,988 (52.1%) | 1,870 (31.3%) |
| AKI cases were determined based on urine output criteria (n(%)) | 1722 (45.1%) | 1486 (24.9%) |
| Age, years (mean (s.d.)) | 60 (18.0) | 60 (18.3) |
| Male gender (n(%)) | 2,169 (56.8%) | 3,404 (57.0%) |
| Comorbidities (n(%)): |  |  |
| Hypertension | 1,625 (42.6%) | 2,430 (40.7%) |
| Diabetes | 527 (13.8%) | 1,253 (21.0%) |
| Congestive heart failure | 593 (15.5%) | 1,172 (19.6%) |
| Chronic pulmonary disease | 550 (14.4%) | 795 (13.3%) |
| Chronic kidney disease | 379 (10.0%) | 395 (6.6%) |
| Chronic liver disease | 201 (5.3%) | 108 (1.8%) |
| **Primary ICD-9 diagnosis (n(%)):** |  |  |
| Sepsis, including pneumonia | 713 (18.7%) | 2,290 (38.3%) |
| Cardiovascular | 1,367 (35.8%) | 1,531 (25.6%) |
| Other Respiratory | 670 (17.6%) | 642 (10.7%) |
| Neurological | 401 (10.5%) | 280 (4.7%) |
| Others | 665 (17.4%) | 1,232 (20.6%) |
| **Ethnicity (n(%)):** |  |  |
| Black | 264 (6.9%) | 420 (7.0%) |
| Hispanic | 66 (1.7%) | 273 (4.6%) |
| Asian | 38 (1.0%) | 202 (3.4%) |
| White | 3,234 (84.8%) | 4,140 (69.3%) |
| Other | 214 (5.6%) | 940 (15.7%) |
| Initial SOFA (mean (s.d.)) | 5.8 (3.3) | 4.1 (3.0) |
| Cardiac surgery (n(%)) | 149 (3.9%) | 122 (2.0%) |
| Mechanical ventilation (n(%)) | 1,276 (33.4%) | 2,830 (47.4%) |
| Vasopressors (n(%)) | 1,006 (26.4%) | 1,666 (27.9%) |
| Need for renal replacement therapy (RRT) (n(%)) | 68 (1.8%) | 57 (0.95%) |
| Length of stay, days (median, (IQR)) | 2.6 (1.6-4.8) | 2.8 (1.8-5.0) |
| ICU mortality rate | 10.00% | 8.60% |

### Model Development and Evaluation in eICU Cohort

Compared with the baseline algorithm (logistic regression algorithm) in eICU cohort, we found that AdaBoost and Gradient Boosting Machine (GBM) achieved higher AUC than other algorithms in predicting task of all four configurations of the time-series models. The model 6-6 configuration in general yielded the highest performance. Using model 6-6, the AUC of logistic regression is 0.8385, while the AUC was higher in the AdaBoost (0.8859) and GBM (0.8522). AUC results of all five-fold cross validation across four different configurations of the time-series models and eight different machine learning algorithms are shown in Figure 4A. Precision and Recall of the models are also demonstrated in Figure S1A and Figure S2A. The model performance (mean values) on eICU cohort testing sets is shown in Table 3. Other performance metrics on eICU cohort testing sets MIMIC-III such as F1 score, negative predictive value (NPV), and specificity are also available at Table S3.

**Table 3.** Comparison of model performance (mean values across five-fold validation results) in eICU internal validation. Abbreviations: AUC: area under the ROC curve. L2 stands for Ridge regularization.

| **Models and algorithms** | Logistic Regression (Baseline) | Decision tree (DT) | AdaBoost | Gradient Boosting Machine (GBM) | Random forest (RF) | Neural Networks (NN) | Neural Networks (NN with L2) | Long short-term memory networks (LSTM) |
| --- | --- | --- | --- | --- | --- | --- | --- | --- |
| **Model 6-6** |  |  |  |  |  |  |  |  |
| AUC | 0.8385 | 0.5937 | **0.8859** | 0.8522 | 0.7954 | 0.7716 | 0.7701 | 0.6154 |
| Precision | 0.6934 | 0.5209 | **0.7381** | 0.5430 | 0.5503 | 0.5328 | 0.5295 | 0.5056 |
| Recall | 0.5353 | 0.5937 | 0.5762 | 0.5307 | 0.5262 | **0.6016** | 0.5975 | 0.5492 |
| **Model 6-12** |  |  |  |  |  |  |  |  |
| AUC | 0.8402 | 0.5856 | **0.8634** | 0.8385 | 0.7509 | 0.7636 | 0.7695 | 0.6192 |
| Precision | 0.7051 | 0.5185 | **0.7073** | 0.5707 | 0.5437 | 0.5377 | 0.5370 | 0.5002 |
| Recall | 0.5380 | 0.5856 | 0.5814 | 0.5551 | 0.5197 | 0.6014 | **0.6040** | 0.5094 |
| **Model 12-6** |  |  |  |  |  |  |  |  |
| AUC | 0.8583 | 0.5962 | **0.8804** | 0.8459 | 0.7766 | 0.7692 | 0.7751 | 0.5727 |
| Precision | 0.7152 | 0.5229 | **0.7339** | 0.5742 | 0.5504 | 0.5269 | 0.5307 | 0.5022 |
| Recall | 0.5524 | 0.5962 | 0.5913 | 0.5584 | 0.5301 | 0.6147 | **0.6262** | 0.5151 |
| **Model 12-12** |  |  |  |  |  |  |  |  |
| AUC | 0.8505 | 0.5820 | **0.8509** | 0.8244 | 0.7175 | 0.7467 | 0.7286 | 0.6100 |
| Precision | 0.6997 | 0.5187 | **0.7412** | 0.5746 | 0.5454 | 0.5249 | 0.5198 | 0.5005 |
| Recall | 0.5443 | 0.5820 | 0.5784 | 0.5496 | 0.5177 | **0.6076** | 0.5931 | 0.5098 |

**Figure 4.**
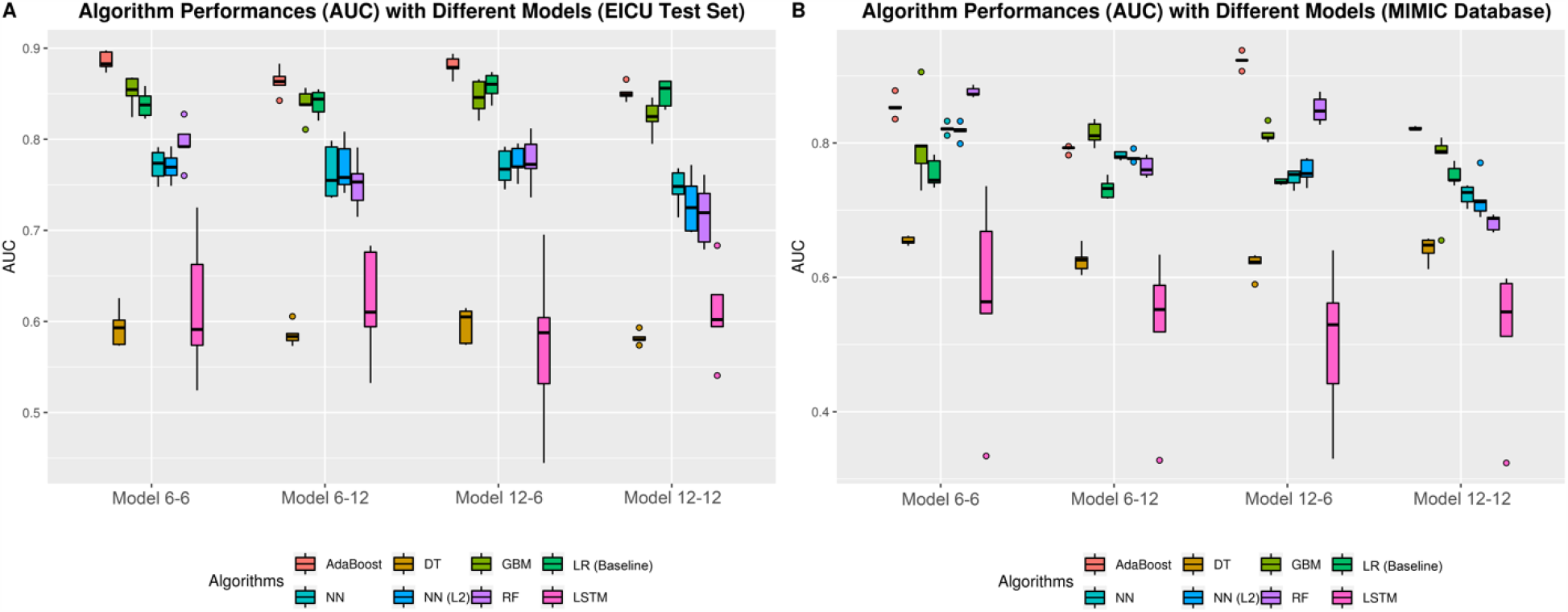
Boxplot of all five-folds recall results across four different configurations of the time-series models (model 6-6, model 6-12, model 12-6, model 12-12) and eight different machine learning algorithms on eICU testing set (A) and external validation database MIMIC-III (B). Abbreviations: DT: Decision Tree; GBM: Gradient Boosting Machine; LR: Logistic Regression; NN: Neural Networks; NN (L2): Neural Networks with L2 regularization; RF: Random Forest; LSTM: Long short-term memory networks.

### Model External Validation and Evaluation in MIMIC-III Cohort

After external validation on four configurations of the time-series models which developed by eight different machine learning algorithms, we found model 12-6 developed by AdaBoost achieved the highest AUC (0.9228) on external validation set. For some models, the higher performance is noted on external validation than model development stage due to the differences between eICU and MIMIC-III cohorts (for example, the onset of AKI in MIMIC-III is more imbalanced), yet the evaluations are comparable within each database cohorts. AUC results of all five-folds results across four different configurations of the time-series models and eight different machine learning algorithms on MIMIC-III external validation set are presented in Figure 4B. The model performances (mean values) on external validation cohort MIMIC-III of AUC, precision, and recall are shown in Table 4. Other performance metrics on external validation cohort MIMIC-III such as F1 score, negative predictive value (NPV), and specificity are also available at Table S4.

**Table 4.** Comparison of model performance (mean values across five-fold validation results) in MIMIC-III cohort for external validation. Abbreviations: AUC: area under the ROC curve. L2 stands for Ridge regularization.

| **Models and algorithms** | Logistic Regression (Baseline) | Decision tree (DT) | AdaBoost | Gradient Boosting Machine (GBM) | Random forest (RF) | Neural Networks (NN) | Neural Networks (NN with L2) | Long short-term memory networks (LSTM) |
| --- | --- | --- | --- | --- | --- | --- | --- | --- |
| **Model 6-6** |  |  |  |  |  |  |  |  |
| AUC | 0.7550 | 0.6544 | 0.8542 | 0.7993 | **0.8760** | 0.8213 | 0.8175 | 0.5696 |
| Precision | 0.5018 | 0.5123 | 0.5103 | 0.5059 | 0.5558 | 0.5073 | 0.5071 | **0.5638** |
| Recall | 0.5395 | 0.6544 | 0.5006 | 0.5648 | 0.5692 | 0.6857 | **0.6886** | 0.5267 |
| **Model 6-12** |  |  |  |  |  |  |  |  |
| AUC | 0.7322 | 0.6253 | 0.7911 | **0.8142** | 0.7642 | 0.7811 | 0.7788 | 0.5241 |
| Precision | 0.5018 | 0.5080 | 0.5039 | 0.5108 | **0.5449** | 0.5064 | 0.5064 | 0.5268 |
| Recall | 0.5387 | 0.6253 | 0.5017 | 0.5987 | 0.6241 | **0.6690** | 0.6647 | 0.5136 |
| **Model 12-6** |  |  |  |  |  |  |  |  |
| AUC | 0.7422 | 0.6192 | **0.9228** | 0.8131 | 0.8502 | 0.7479 | 0.7579 | 0.5006 |
| Precision | 0.5019 | 0.5077 | 0.5044 | 0.5076 | **0.5593** | 0.5047 | 0.5051 | 0.5343 |
| Recall | 0.5412 | 0.6192 | 0.5014 | 0.6005 | 0.6291 | 0.6695 | **0.6773** | 0.5077 |
| **Model 12-12** |  |  |  |  |  |  |  |  |
| AUC | 0.7522 | 0.6419 | **0.8215** | 0.7662 | 0.6816 | 0.7224 | 0.7173 | 0.5147 |
| Precision | 0.5022 | 0.5095 | 0.5038 | 0.5066 | 0.5213 | 0.5044 | 0.5043 | **0.5250** |
| Recall | 0.5451 | 0.6419 | 0.5011 | 0.5792 | 0.6223 | **0.6512** | 0.6459 | 0.5140 |

### Secondary Outcomes

Secondary outcomes of eICU and MIMIC-III databases are summarized in Table 5. P-value is derived according to alternative hypothesis: true Spearman *ρ* is not equal to 0 when comparing the variable with the onset of AKI. The patients who were diagnosed with AKI within the first week of ICU were associated with higher ICU mortality (eICU: 261 (13.1%) vs 121 (6.6%), spearman *ρ* = 0.1083, p<0.01; MIMIC-III: 284 (15.2%) vs 230 (5.6%), spearman *ρ* = 0.1585, p<0.01), renal replacement therapy (RRT) (eICU: 63 (3.2%) vs 5 (0.3%), spearman *ρ* = 0.1093, p<0.01; MIMIC-III: 47 (2.5%) vs 10 (0.2%), spearman *ρ* = 0.1083, p<0.01), longer ICU length of stay (eICU: median=3.1 (2.2-3.9) vs median=2.0 (1.1-2.5), spearman *ρ* = 0.2738, p<0.01; MIMIC-III: median=4.8 (IQR: 3.5-5.8) vs median=2.2 (IQR: 1.2-2.6), spearman *ρ* = 0.4185, p<0.01), and larger ISOFA score (eICU: mean=6.2 (standard deviation=3.4) vs mean=5.3 (standard deviation=3.1), spearman *ρ* = 0.1229, p<0.01; MIMIC-III: mean=4.9 (standard deviation=3.3) vs mean=3.8 (standard deviation=2.8), spearman *ρ* = 0.1623, p<0.01).

**Table 5.**
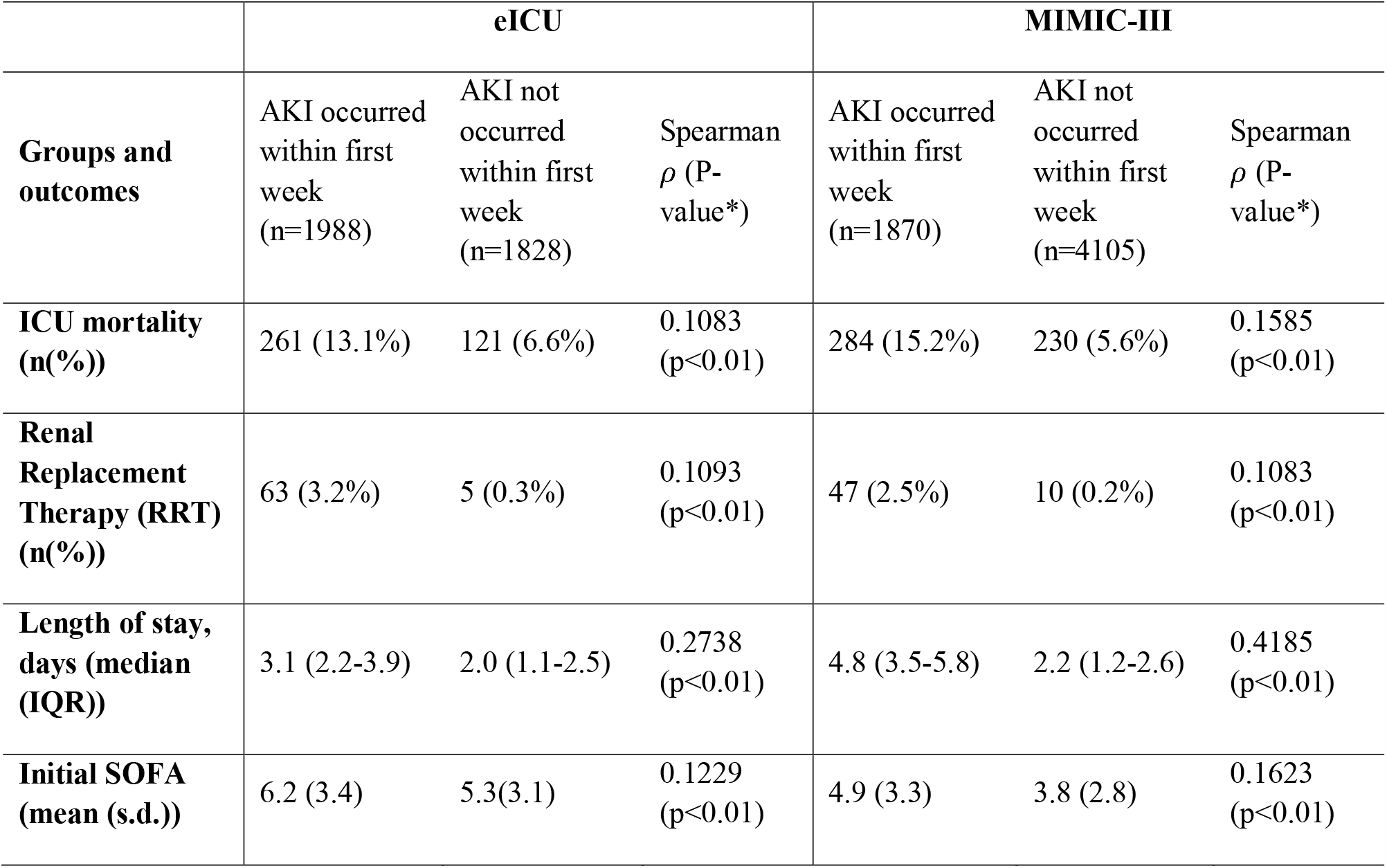
Secondary outcomes of eICU and MIMIC-III databases. s.d.: standard deviation in short. IQR: interquartile range in short. *P-value is derived according to alternative hypothesis: true Spearman *ρ* is not equal to 0 when comparing the variable with the onset of AKI.

|  | **eICU** |  |  | **MIMIC-III** |  |  |
| --- | --- | --- | --- | --- | --- | --- |
| **Groups and outcomes** | AKI occurred within first week (n=1988) | AKI not occurred within first week (n=1828) | Spearman $\rho$ (P-value*) | AKI occurred within first week (n=1870) | AKI not occurred within first week (n=4105) | Spearman $\rho$ (P-value*) |
| **ICU mortality (n(%))** | 261 (13.1%) | 121 (6.6%) | 0.1083 (p<0.01) | 284 (15.2%) | 230 (5.6%) | 0.1585 (p<0.01) |
| **Renal Replacement Therapy (RRT) (n(%))** | 63 (3.2%) | 5 (0.3%) | 0.1093 (p<0.01) | 47 (2.5%) | 10 (0.2%) | 0.1083 (p<0.01) |
| **Length of stay, days (median (IQR))** | 3.1 (2.2-3.9) | 2.0 (1.1-2.5) | 0.2738 (p<0.01) | 4.8 (3.5-5.8) | 2.2 (1.2-2.6) | 0.4185 (p<0.01) |
| **Initial SOFA (mean (s.d.))** | 6.2 (3.4) | 5.3(3.1) | 0.1229 (p<0.01) | 4.9 (3.3) | 3.8 (2.8) | 0.1623 (p<0.01) |

### Relationships between Prediction Performances and Different Time-Series Models

As eight different algorithms with five-fold cross-validation produces 40 results per each time series model (model 6-6, model 6-12, model 12-6, model 12-12), violin plot (Figure 5) and pairwise paired t-test (Table 6) are used to compare the overall performances across time series models. In figure 5, we demonstrated that given fixed feature collecting window, models with closer gap tend to have better AUC in average and in pairwise paired t-test (model 6-6: 0.7653 > model 6-12: 0.7539, p-value: 0.0238; model 12-6: 0.7593 > model 6-12: 0.7388, p-value: 0.0218). Moreover, with fixed gap window, models with shorter feature collecting window also tend to have better AUC in average and in pairwise paired t-test (model 6-6: 0.7653 > model 12-6: 0.7593, p-value: 0.459; model 6-12: 0.7539 > model 12-12: 0.7388, p-value: 0.0227), Such findings suggest that the prediction performance improved with closer gaps, while increasing the lag of time-series data doesn’t improve the model performances.

**Table 6.** Pairwise paired t-test comparison among the five-fold cross-validation AUC results over four different time-series models.

|  |  |
| --- | --- |
|  | Set 2 |

|  |  | Model 6-6 |  | Model 6-12 |  | Model 12-6 |  | Model 12-12 |  |
| --- | --- | --- | --- | --- | --- | --- | --- | --- | --- |
|  |  | t | P | t | P | t | P | t | P |
| Set<br>1 | Model 6-6 | - | - | 2.35 | 2.38E-02* | 0.75 | 4.59E-01 | 4.22 | 1.40E-04* |
|  | Model 6-12 | -2.35 | 2.38E-02* | - | - | -0.73 | 4.71E-01 | 2.37 | 2.27E-02* |
|  | Model 12-6 | -0.75 | 4.59E-01 | 0.73 | 4.71E-01 | - | - | 2.39 | 2.18E-02* |
|  | Model 12-12 | -4.22 | 1.40E-04* | -2.37 | 2.27E-02* | -2.39 | 2.18E-02* | - | - |
| Notes: t denotes the pairwise paired Student's t-test statistic, P denotes the p-value obtained. P-value < 0.05 are significant and indicated with * symbol. |  |  |  |  |  |  |  |  |  |

**Figure 5.**
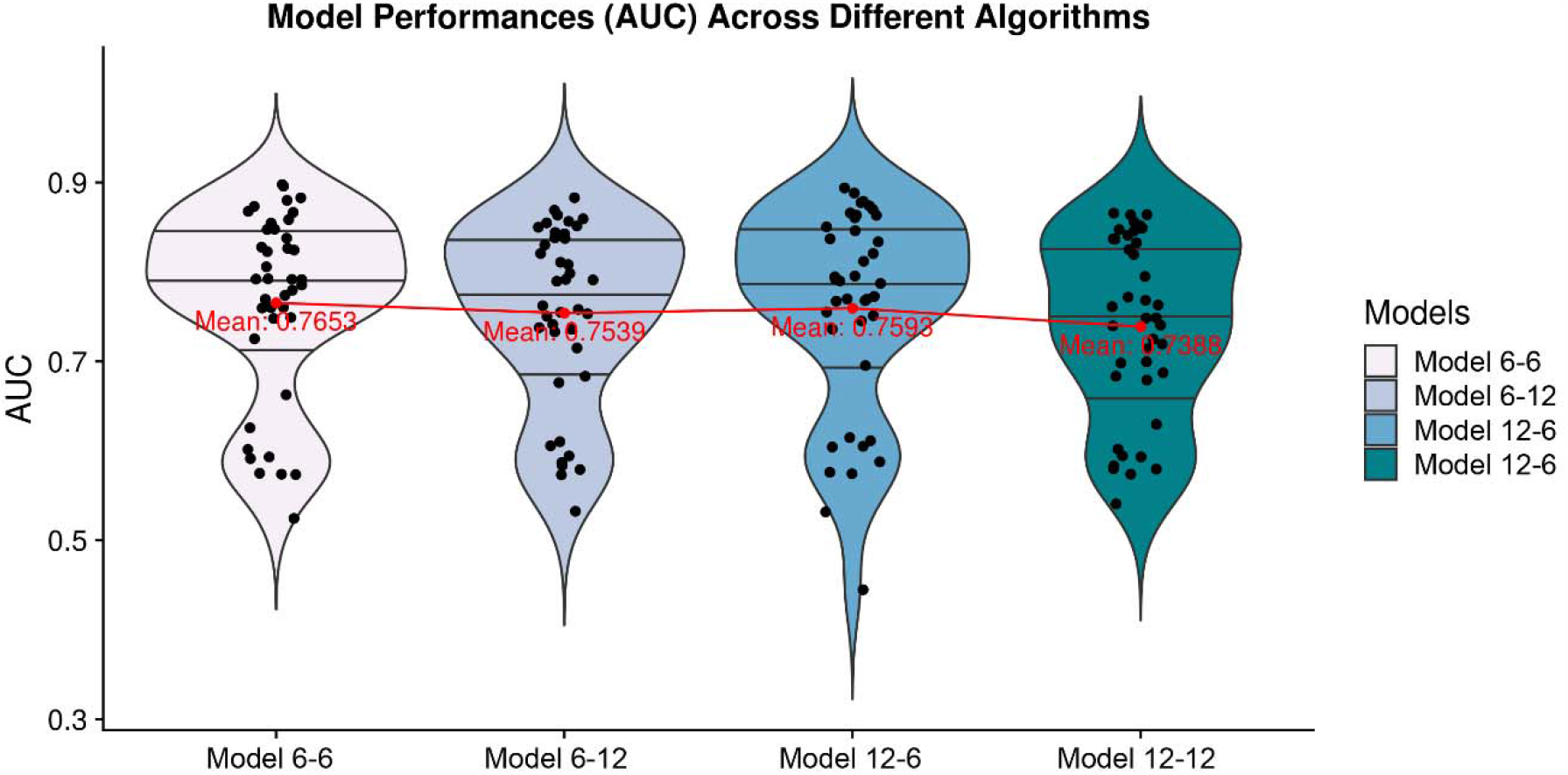
Model performances (AUC) violin plot of four different configurations of the models (model 6-6, model 6-12, model 12-6, model 12-12) across eight different algorithms with five-fold cross-validation. Black solid horizontal lines in each violin plot are the interquartile range lines. Red points and lines indicate the mean performances (AUC) over four models.

### Identifying Feature Importance

The feature importance identified by the AdaBoost algorithm is determined by the average feature importance over all ensembled trees. We categorized eight groups of features and analyzed feature importance by Gini importance (mean decrease impurity). The list of categorized groups is shown in Table 1.

To gain insights into the relevance of each feature, Figure 6 summarized and ranked the most critical variables in model 6-6 based on the averaged Gini feature importance using AdaBoost algorithm in model development (eICU database). Note that vitals, labs, topics and static data are important for the prediction task, and the most important group of features was vital signs. Specifically, fluid balance-related features (OUTPUT_12HR, OUTPUT_6HR, INPUT_12HR, OUTPUT_24HR), AST, AG, SCr, UN, CA_ION, K_ION in the class of laboratory values, SYS_BP, RR, HR in the class of vital signs, HOURS in the class of others, and WEIGHT in the class of Demographic, are the most important predictors for the early AKI prediction.

**Figure 6.**
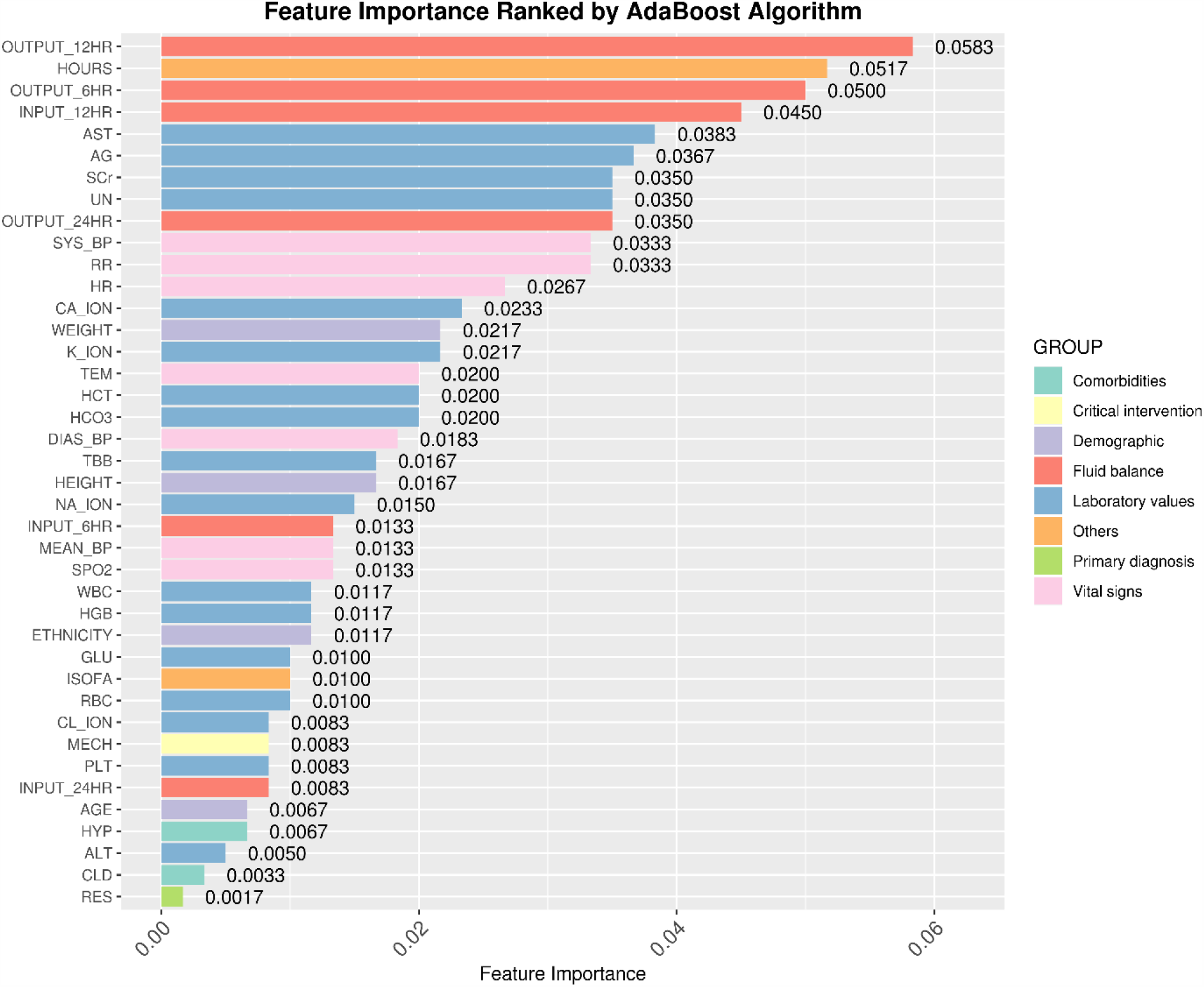
Identified top important features in eICU database. Values and rankings are based on time-series model 6-6 and the averaged Gini importance from AdaBoost algorithm. Features are colored group-wisely according to the categories listed in Table 1. The higher value indicates the higher significance of the feature.

## Discussion

Electronic healthcare systems (EHR) provides real-world clinical data not only for secondary analysis, but also for developing the artificial intelligence-based platform to assist clinicians to identify potential critical events, such as the onset of AKI ^82-84^. Since there is no effective treatment of AKI, the prevention of AKI becomes more critical ^2, 59, 85-89^, thus an early warning system which may reduce the risk of exacerbating injury is necessary and in urgent demand.

Though some studies on early warning system have been successfully reduced the nephrotoxic medication in AKI and prevented the contrast-induced AKI ^90-92^, several other studies showed the failure of those methods ^93-95^, which mainly due to the lack of external validation. Therefore, the study with external validation are necessary before any developed method be applied to the clinical setting, especially the externally validated prediction models are relatively rare ^19, 42, 56^.

In this paper, we have integrated most of the routinely available ICU data based on two large ICU databases with general population to developed and externally validated an ideal artificial intelligence assisted early warning system for predicting the onset of AKI within the first week of ICU stay. Our workflow and time-series models addressed several topics that have been discussing over these days. For example, we used both oliguric and SCr as diagnostic criteria of the KDIGO to identify the onset of AKI and included most of physiological and laboratory parameters as predictors, which contained different clinical commonly used urine output trends. As a result, we have got an ideal incidence of AKI in both eICU and MIMIC-III cohort, which would minimize the potential mistakes and maximize the accuracy and improving of predicting model performance in clinical situation. Moreover, we fully made use of all routinely available ICU and clinical features as predictors, to optimize and improve the model performances.

Using multiple types of ICU and clinical features to predict AKI, a complex clinical syndrome, is very challenging and difficult as any feature or small changes may cause great impact on outcomes. Machine learning technique has been successfully applied to predict critical events under a fast-paced, data-overloaded setting of ICU, which could provide new insights into evidence-based decision support ^96-101^. Different machine learning algorithms usually used to capture the real relationships across different data types which may provide advantages for predicting AKI far in advance of onset with high sensitivity and specificity. As previous studies, many machine learning algorithms have been used training many ideal models in predicting AKI, such as random forest, multivariate regression model, boosted ensembles of decision trees, etc. ^26, 31, 59^. Based on these previous studies, we take a further step by fully utilizing two independent databases and applied eight machine learning algorithms with 52 comprehensive features of different data types for a large-scale model assessments and analyses.

Our current study has several strengths. First, we integrated most of the routinely available ICU data, including 52 features under different data types, which would be considered containing the largest number of predictors. Second, we not only used both urine output and SCr as diagnostic criteria to identify the onset of AKI, but also included physiological (especially urine output) and laboratory trends as predictors to improve model performance. Third, we compared 8 modern machine learning algorithms for further analysis to select the best model of predicting task. Fourth, we developed and externally validated an ideal model based on two independent general population ICU databases. Last but not least, we did not only rely on using data at a single time point (admission to hospital), but also combining static and time-varying variables as a time series dataset to achieve accurate prediction model performance.

From the results and performances across four time-series models and various machine learning algorithms, we proved the feasibility of deploying artificial intelligence assisted early warning system for AKI prediction. We also showed that AdaBoost, one of the ensemble machine learning algorithms, could be a desired model to predict the onset of AKI. The performances measured by AUC is decent and could be applied in real-time to assist clinicians’ decision in the future. While the across-model performances demonstrated that increasing the lag (feature collecting window), or in another word, how many hours we look back, doesn’t help improve the performance, justified that model 6-6 with 6 hours feature collecting window is enough for the future AKI inference.

For feature importance ranking of AKI prediction, the top 15 features are critical and closely related to AKI development. We justified and validated that the fluid balance and laboratory values are not only the AKI criteria of the KDIGO, but also directly associated with the kidney function (such as OUTPUT_12HR, OUTPUT_6HR, INPUT_12HR, SCr, UN, CA_ ION, K_ION, etc.) ^54^. Most vital signs, which are used to assess patients’ physiological stability, are important for prediction as the AKI prediction should be in real-time. Hours stayed in ICU and weight are also indicated as the most important features which have already been reported that both of them have a great association with AKI ^102^. The model developed has the potential capability of serving as an “alert” for early warning of AKI, which would make the electronic healthcare systems more artificially intelligence and enable bedside application.

Although our study bridged many gaps in AKI forecasting research, this study may still exist some inevitable limitations to be addressed in the future. First, the AKI is defined with the baseline creatinine chosen as first in stay, which could miss the true incidence of AKI by up to 30% who already had AKI at that time ^103^. However, as we face the limitation of the data, the safest way for now is to exclude that group of patients with unclear information. Same exclusion due to the data limitation also happened in the withdrawal of treatment, which contains patients who did not receive mechanical ventilation and died during ICU stay. Second, we predict the onset of AKI at any stage, without predicting the precise stage. In the future, multi-class classification or even regression might be considered instead of binary classification. Third, comorbidities were included as “static” variables, since they could not be accurately timestamped due to the limited information of the eICU and MIMIC-III databases. Fourth, the predictors may need more effort on improving the recall score – with well-performed AUC and precision, the recall is not so promising due to some positive samples (AKI) are misclassified. Fifth, although we used two non-overlapping general population ICU databases, both are from the United States hospitals. Thus, cross-national and multi-background validations are necessary. Sixth, we only evaluated two feature windows (6 and 12 hours), two gap windows (6 and 12 hours) and one prediction window (1 hour) due to the data and computational limitation. In the future, we may put more efforts on various time-series models. Last but not the least, study design in this work relied on retrospective data investigation which may cause missing some important information than a prospective study and could not have any result about the impact analysis between model prediction and patients’ outcomes. Therefore, the model also requires further external validation based on different background populations and may only be used inside the research arena.

## Conclusion

Recent reviews and comments showed that the prediction of AKI in the ICU is relatively difficult and with several limitations. To answer these limitations and problems, We have developed an artificial intelligence assisted early warning model for predicting the onset of AKI within the first week of ICU stay, which identified by using both oliguric and SCr diagnostic criteria in multi-center eICU database and externally validated in single-center MIMIC-III database. We integrated most routinely available ICU data and demonstrated the model with 6 hours feature and 6 hours gap developed by AdaBoost achieved optimal performances measured with AUC, precision, and recall among 8 prevalent machine learning algorithms.

## Data Availability

Datasets are used from the collection of eICU® Collaborative Research Database (eICU, https://eicu-crd.mit.edu/) (v1.2) and Medical Information Mart for Intensive Care III (MIMIC-III, https://mimic.physionet.org/) (v1.4).

https://mimic.physionet.org/

## Data Availability Statement

Datasets are used from the publically available collection of eICU® Collaborative Research Database (eICU, https://eicu-crd.mit.edu/) (v1.2) and Medical Information Mart for Intensive Care III (MIMIC-III, https://mimic.physionet.org/) (v1.4).

## Code Availability Statement

Experiments were conducted with two open-source libraries: the machine learning library scikit-learn (https://scikit-learn.org/stable/), PyTorch (https://pytorch.org/), which provides the public implementations of all listed algorithms. Libraries were called from python 3.7 environment (https://www.python.org/).

## Competing interests

The authors declare no conflict of interests.

## Table Legends

Table S1. eICU and MIMIC-III databases exclusion criteria.

Table S2. Most commonly missing features according to all timestamps in eICU database. Total missing count was the count of total missing occurrence of the associated feature. Missing ratio was the ratio of missing observations to the entire observations (427,527 timestamps in total). All abbreviations are referred to eICU and MIMIC-III databases. Features with high missing ratio were dropped.

Table S3. Comparison of other performance metrics (mean values across five-fold validation results) on eICU internal validation. Abbreviations: NPV: negative predictive value. L2 stands for Ridge regularization.

Table S4. Comparison of other performance metrics (mean values across five-fold validation results) on MIMIC-III cohort for external validation. Abbreviations: NPV: negative predictive value. L2 stands for Ridge regularization.

## Notes

**Conflicts of Interest:** The authors declare that they have no conflict of interest.

### Competing Interest Statement

The authors have declared no competing interest.

### Funding Statement

There is no financial funding or interest to report.

### Summary of Updates

Author list and author contribution corrected.

