## Supplemental tables for "Artificial Intelligence Assisted Early Warning System for Acute Kidney Injury Driven by Multi-Center ICU Database": Table S1.docx

| **Feature names** | **Unit** | **Exclusion criteria** |
| --- | --- | --- |
| Age |  | > 200 |
| Heart rate | /min | > 300 |
| Systolic blood pressure | mmHg | > 400 |
| Diastolic blood pressure | mmHg | > 300 |
| Mean blood pressure | mmHg | > 300 |
| Respiratory rate | /min | > 70 |
| Temperature | [℃](http://www.jiaocheng8.com/word/503.html) | < 10 or > 50 |
| Oxygen saturation | % | > 100 |
| FiO2 | % | > 100 or < 20 |
| Sodium | mmol/L | > 200 |
| Potassium | mmol/L | > 10000 |
| Chloride | mmol/L | > 10000 |
| Anion_gap | mmol/L | > 10000 |
| Glucose | mg/dL | > 10000 |
| Bicarbonate | mmol/L | > 10000 |
| White blood cell count | K/uL | > 1000 |
| Hemoglobin | g/dL | > 50 |
| Hematocrit | % | > 100 |
| Platelets | K/uL | > 10000 |
| Totalbilirubin | mg/dL | > 150 |
| Blood urea nitrogen | g/dL | > 300 |
| Blood creatinine | mg/dL | > 150 |

Table S1. eICU and MIMIC-III databases exclusion criteria.
