## Supplemental tables for "Artificial Intelligence Assisted Early Warning System for Acute Kidney Injury Driven by Multi-Center ICU Database": Table S2.docx

| **Feature names** | **Total missing count** | **Missing ratio** |
| --- | --- | --- |
| PH | 298,181 | 0.6975 |
| LAC | 251,583 | 0.5885 |
| APTT | 187,918 | 0.4395 |
| P_ION | 183,157 | 0.4284 |
| INR | 126,116 | 0.2950 |
| TOTALBILIRUBIN | 119,355 | 0.2792 |
| ALT | 118,531 | 0.2772 |
| ALB | 116,456 | 0.2724 |
| AST | 116,029 | 0.2714 |
| MG_ION | 87,900 | 0.2056 |
| RDW | 68,596 | 0.1604 |
| MEAN_BP | 35,772 | 0.0837 |
| SPO2 | 33,095 | 0.0774 |
| HR | 32,625 | 0.0763 |
| TEM | 31,689 | 0.0741 |
| RESPRATE | 30,968 | 0.0724 |
| DIAS_BP | 30,796 | 0.0720 |
| SYS_BP | 30,796 | 0.0720 |
| RBC | 11,438 | 0.0268 |
| HEMOGLOBIN | 9,463 | 0.0221 |
| WBC | 9,005 | 0.0211 |
| CA_ION | 8,102 | 0.0190 |
| PLT | 8,079 | 0.0189 |
| HEMATOCRIT | 7,765 | 0.0182 |
| UN | 6,124 | 0.0143 |
| HCO3 | 5,635 | 0.0132 |
| ANION_GAP | 5,635 | 0.0132 |
| SCr | 5,593 | 0.0131 |
| CL_ION | 5,502 | 0.0129 |
| SODIUM | 5,138 | 0.0120 |
| K_ION | 5,086 | 0.0119 |
| GLU | 3,360 | 0.0079 |
| ETHNICITY | 541 | 0.0013 |
| GENDER | 141 | 0.0003 |
| BMI | 99 | 0.0002 |

Table S2. Most commonly missing features according to all timestamps in eICU database. Total missing count was the count of total missing occurrence of the associated feature. Missing ratio was the ratio of missing observations to the entire observations (427,527 timestamps in total). All abbreviations are referred to eICU and MIMIC-III databases. Features with high missing ratio were dropped.
