## Supplemental tables for "Artificial Intelligence Assisted Early Warning System for Acute Kidney Injury Driven by Multi-Center ICU Database": Table S3.docx

| **Models and algorithms** | Logistic Regression (Baseline) | Decision tree (DT) | AdaBoost | Gradient Boosting Machine (GBM) | Random forest (RF) | Neural Networks (NN) | Neural Networks (NN with L2) | Long short term-memory networks (LSTM) |
| --- | --- | --- | --- | --- | --- | --- | --- | --- |
| **Model 6-6** |  |  |  |  |  |  |  |  |
| F1-score | 0.5570 | 0.5226 | **0.6134** | 0.5337 | 0.5325 | 0.5423 | 0.5377 | 0.5072 |
| NPV | 0.9854 | 0.9874 | 0.9866 | 0.9852 | 0.9851 | **0.9876** | 0.9875 | 0.9851 |
| specificity | **0.9982** | 0.9251 | 0.9973 | 0.9870 | 0.9900 | 0.9463 | 0.9441 | 0.9686 |
| **Model 6-12** |  |  |  |  |  |  |  |  |
| F1-score | 0.5610 | 0.5174 | **0.6149** | 0.5595 | 0.5247 | 0.5470 | 0.5481 | 0.5009 |
| NPV | 0.9841 | 0.9859 | 0.9855 | 0.9847 | 0.9834 | 0.9864 | **0.9865** | 0.9832 |
| specificity | **0.9980** | 0.9160 | 0.9960 | 0.9862 | 0.9882 | 0.9463 | 0.9487 | 0.9642 |
| **Model 12-6** |  |  |  |  |  |  |  |  |
| F1-score | 0.5817 | 0.5254 | **0.6292** | 0.5636 | 0.5354 | 0.5307 | 0.5369 | 0.5034 |
| NPV | 0.9845 | 0.9863 | 0.9859 | 0.9848 | 0.9838 | 0.9870 | **0.9874** | 0.9830 |
| specificity | **0.9976** | 0.9234 | 0.9963 | 0.9870 | 0.9868 | 0.9198 | 0.9233 | 0.9837 |
| **Model 12-12** |  |  |  |  |  |  |  |  |
| F1-score | 0.5696 | 0.5169 | **0.6161** | 0.5573 | 0.5221 | 0.5260 | 0.5132 | 0.5005 |
| NPV | 0.9826 | 0.9843 | 0.9839 | 0.9828 | 0.9816 | **0.9854** | 0.9849 | 0.9815 |
| specificity | **0.9974** | 0.9114 | 0.9969 | 0.9870 | 0.9857 | 0.9105 | 0.8914 | 0.9579 |

Table S3. Comparison of other performance metrics (mean values across five-fold validation results) on eICU internal validation. Abbreviations: NPV: negative predictive value. L2 stands for Ridge regularization.
