## Supplemental tables for "Artificial Intelligence Assisted Early Warning System for Acute Kidney Injury Driven by Multi-Center ICU Database": Table S4.docx

| **Models and algorithms** | Logistic Regression (Baseline) | Decision tree (DT) | AdaBoost | Gradient Boosting Machine (GBM) | Random forest (RF) | Neural Networks (NN) | Neural Networks (NN with L2) | Long short term-memory networks (LSTM) |
| --- | --- | --- | --- | --- | --- | --- | --- | --- |
| **Model 6-6** |  |  |  |  |  |  |  |  |
| F1-score | 0.4670 | 0.5048 | 0.4999 | 0.4817 | **0.5598** | 0.4661 | 0.4625 | 0.4658 |
| NPV | 0.9946 | 0.9960 | 0.9940 | 0.9949 | 0.9949 | 0.9967 | **0.9968** | 0.9943 |
| specificity | 0.8497 | 0.9201 | **0.9996** | 0.8793 | 0.9932 | 0.8166 | 0.8060 | 0.8479 |
| **Model 6-12** |  |  |  |  |  |  |  |  |
| F1-score | 0.4644 | 0.4892 | 0.5018 | 0.5045 | **0.5635** | 0.4551 | 0.4562 | 0.4344 |
| NPV | 0.9941 | 0.9954 | 0.9936 | 0.9949 | 0.9952 | **0.9963** | 0.9962 | 0.9939 |
| specificity | 0.8398 | 0.8891 | **0.9973** | 0.9289 | 0.9826 | 0.7853 | 0.7889 | 0.7536 |
| **Model 12-6** |  |  |  |  |  |  |  |  |
| F1-score | 0.4626 | 0.4886 | 0.5014 | 0.4905 | **0.5789** | 0.3984 | 0.4073 | 0.4583 |
| NPV | 0.9942 | 0.9953 | 0.9936 | 0.9950 | 0.9952 | **0.9970** | **0.9970** | 0.9937 |
| specificity | 0.8334 | 0.8893 | **0.9973** | 0.8969 | 0.9864 | 0.6306 | 0.6536 | 0.8330 |
| **Model 12-12** |  |  |  |  |  |  |  |  |
| F1-score | 0.4607 | 0.4879 | 0.5007 | 0.4832 | **0.5286** | 0.3794 | 0.3813 | 0.4339 |
| NPV | 0.9937 | 0.9952 | 0.9929 | 0.9942 | 0.9947 | **0.9966** | 0.9964 | 0.9931 |
| specificity | 0.8240 | 0.8750 | **0.9984** | 0.8774 | 0.9571 | 0.5824 | 0.5884 | 0.7514 |

Table S4. Comparison of other performance metrics (mean values across five-fold validation results) on MIMIC-III cohort for external validation. Abbreviations: NPV: negative predictive value. L2 stands for Ridge regularization.
